# Atrial fibrillation in adult congenital heart disease in Asia

**DOI:** 10.1101/2023.04.20.23288895

**Authors:** Shuenn-Nan Chiu, Wei-Chieh Tseng, Chun-Wei Lu, Ming-Tai Lin, Chun-An Chen, Jou-Kou Wang, Mei-Hwan Wu

**Author notes:** Address for correspondence: Shuenn-Nan Chiu, Department of Pediatrics, National Taiwan University Hospital, No. 7, Chung-Shan South Road, Taipei, Taiwan 100, [ ] #71710. **Disclosure:** The authors have no financial relationships relevant to this article to disclose.

## Abstract

**Introduction:** With the improvement of long-term survival of patients with congenital heart disease (CHD), complications such as atrial fibrillation (AF) have become a concern. This study aimed to determine the epidemiology data and risk factors of AF in adult CHD (ACHD) patients and evaluate the impact of AF on late outcomes using a large ACHD cohort in Asia.

**Method:** This study enrolled all CHD patients older than 18 years of age diagnosed with CHD at National Taiwan University Hospital between 2007 and 2018. Data on patients’ clinical characteristics, electrocardiogram, Holter reports, and follow-up information were collected. AF status was classified as sustained AF, paroxysmal AF, or intra-atrial reentry tachycardia (IART). CHD was categorized as simple, severe, or complex CHD (single ventricle). Primary endpoint was defined as cerebrovascular accidents (CVA) or death.

**Result:** The study included 4403 patients (55.9% women), with 16.4% having severe and 2.9% having complex CHD. The cumulative incidence of AF was 6.9% (54.8% paroxysmal AF, 26.9% sustained AF, and 18.4% IART), which is comparable to the Western countries. The incidence increased with age and was higher in patients with pulmonary hypertension (PH, 27%), complex CHD (12.7%), and metabolic syndrome. The mean onset age of IART, paroxysmal, and sustained AF was 35.7±15.8, 48.4±19.3, and 56.9±14.2 years, respectively. Multivariate Cox regression analysis revealed that male sex, PH, and severe and complex CHD were the most critical risk factors for AF (odds ratio 1.67, 1.91, 3.55, and 12.6, respectively). The 70-year CVA-free survival rate was 67.1% in patients with AF (vs. 80.5% in those without AF, p<0.001). However, multivariate Cox regression analysis identified male sex, PH, severe and complex CHD, and genetic syndrome as the most significant risk factors of the primary endpoint (odds ratio 1.76, 3.38, 2.62 and 19.3, and 8.91, respectively).

**Conclusions:** This large ACHD cohort showed a high cumulative incidence of AF, similar to the Western countries, which increased with age, PH, and CHD severity. CVA-free survival was more closely associated with these factors than with AF.

## Introduction

With advances in perioperative care, the long-term outcome for patients with congenital heart disease (CHD) has significantly improved in recent decades. As a result, the majority of CHD patients can now expect to have a near-normal life expectancy^1^. However, long-term morbidies, such as atrial fibrillation (AF) and its associated complications, have become a major concern. In an earlier study by Bouchardy et al, the projected risk of developing atrial arrhythmia by the age of 65 was close to 50%. ^2^. In a large cohort study of adult CHD (ACHD) patients in Canada, 15% of patients experienced atrial arrhythmia^2^. Similarly, a Swedish nationwide register study found that by the age of 42, 8.3% of all CHD patients had been diagnosed with AF.^3^. Because atrial tachyarrhythmia, particularly AF, is a well-known risk factor for heart failure, cardiovascular events, and death, it is crucial to identify and manage AF in CHD patients as early as possible. The importance of early detection and management of AF in CHD patients cannot be overstated.

In the general population of Western countries, the incidence rate of AF varies from 3.3 to 19.2 per 1,000 person-years, depending on the age of the enrolled population.^4–6^. However, in a recent nationwide population study in Taiwan, the incidence rate was found to be 1.51 per 1,000 person-years^7^, which is comparable to the rate of 1.82 per 1,000 person-years reported in a Korean study^8^. These findings suggest a possible ethnic difference in the incidence of AF, although it is important to note that underestimation, lower cardiovascular risk, and shorter life expectancy may also be contributing factors.^7^.

In a previous study conducted at a large single tertiary center, we found that the presence of pulmonary hypertension (PH) is a significant risk factor for all-cause mortality in patients with CHD, in addition to disease complexity, age, and sex^9^.

Previous studies have also shown that PH is associated with supraventricular arrhythmia, with an incidence as high as 25% in the idiopathic group 1 PH^10^. However, it is currently unknown whether the presence of PH affects the development of AF in ACHD patients. Therefore, in this study, we aimed to investigate the incidence rate, risk factors, and outcomes of AF in a large cohort of Asian ACHD patients, and to explore the association between AF and PH using a comprehensive ACHD database.

## Methods

### Data acquisition

This retrospective study aimed to evaluate atrial fibrillation (AF) in patients with adult congenital heart disease (ACHD) utilizing healthcare records from the integrated medical database at National Taiwan University Hospital (NTUH-iMD), which is the largest ACHD center in Taiwan. The study retrieved clinical, therapeutic, and laboratory data in digitalized format from outpatient, inpatient, and emergency departments. Furthermore, to determine the survival rate and causes of death, the study accessed the death records from the Ministry of Health and Welfare in Taiwan. The study was approved by the Institutional Research Board of the hospital, and due to the anonymity of the data, informed patient consent was waived.

### Patient cohort

This study enrolled adult patients, aged over 18 years, who were diagnosed with CHD between January 2007 and July 2018. Patients who had pre-existing AF at their initial visit to our hospital were excluded from the study. The diagnosis of CHD was based on the International Classification of Diseases, Ninth Revision, Clinical Modification (ICD-9-CM) codes 745.0-747.42 or Tenth Revision ICD-10-CM codes Q20-26. Subsequently, the participants were classified into three subtypes of CHD, namely simple, severe, or complex, as per the previously assigned CHD subtypes (supplementary table 1) ^11, 12^. The complex CHD subgroup included single-ventricle diseases (SV). Common forms of CHD, such as ASD, VSD, endocardial cushion defect (ECD), patent ductus arteriosus (PDA), tetralogy of Fallot (TOF), transposition of the great arteries (TGA), pulmonary atresia with VSD (PA-VSD), and SV were also analyzed separately. SV was defined as the subgroup of patients requiring Fontan type operation.

### Definitions

The categorization of atrial fibrillation (AF) was determined in accordance with the 2014 American Heart Association/American College of Cardiology/Heart Rhythm Society and 2016 European Society of Cardiology guidelines for AF management.

Intra-atrial reentry tachycardia (IART) was defined as cavotricuspid isthmus-dependent atrial flutter or intra-atrial reentry atrial flutter, confirmed by electrocardiogram or electrophysiology study. Paroxysmal AF was defined as self-terminating AF, while sustained AF (persistent or permanent AF) was defined as AF that persists for more than 7 days. The investigators reviewed both the electrocardiogram (ECG) and Holter report to establish the diagnosis of AF. Pulmonary hypertension (PH) associated with CHD and biventricular physiology was defined as a mean pulmonary artery pressure (mPAP) greater than 25 mmHg and pulmonary vascular resistance (PVR) greater than 3 Wood units, as determined by cardiac catheterization, or a peak tricuspid regurgitation (TR) velocity greater than 3.4 m/s (pressure gradient [PG] greater than 46 mmHg) or a TR velocity greater than 2.8 m/s (PG greater than 31 mmHg) with signs of right heart pressure overload by echocardiography^11^. In patients with CHD and single-ventricle physiology, PH was defined as an mPAP equal to or greater than 15 mmHg during admission catheterization, in accordance with previous reports and current guidelines^13, 14^. The primary endpoint of this study was defined as death or cerebrovascular accident (CVA).

### Statistical analysis

All measurements were reported as mean± standard deviation values, unless otherwise specified. For comparisons between groups, numerical data were analyzed using Student’s t-test, and categorical data were analyzed using the Chi-square test. Survival analyses were conducted using the Kaplan-Meier method with log-rank analysis and Cox regression. The CVA-free survival was evaluated from birth to the time of CVA occurrence or death or end of study, with patients censored at loss to follow-up. For survival and Cox regression analysis of AF, follow-up was from birth to the time of AF diagnosis or end of study, with patients censored at death or loss to follow-up. All testing was driven by standard software (SAS v9.4 for Windows; SAS Institute, Cary, NC, USA), setting significance at *p*<0.01.

## Result

### Baseline Characteristics of ACHD Patients

A total of 4,403 ACHD patients were enrolled in this study, of whom 55.9% were women, and the mean age at the last follow-up was 35.4 ± 15.3 years. Of the CHD cases, 16.4% were categorized as severe CHD, and 2.9% as complex CHD. The detailed percentage of specific disease categories were shown in supplementary table 1. The most common forms of simple CHD were ASD, VSD, and PDA, while severe CHD was dominated by TOF and TGA, and complex CHD by heterotaxy.

Baseline demographic data were summarized in Table 1. AF was present in 6.9% of the patients, and PH was observed in 6.9% of the patients with the most common type being PH after defect correction (51.3%). Genetic syndromes accounted for 2.3% of the patients, and 78.9% had a history of previous interventions. The prevalence of hypertension was noted in 8.5%, diabetes mellitus was found in 4.6%, and CVA occurred in 1.5% of the patients in this ACHD cohort.

**Table 1.**
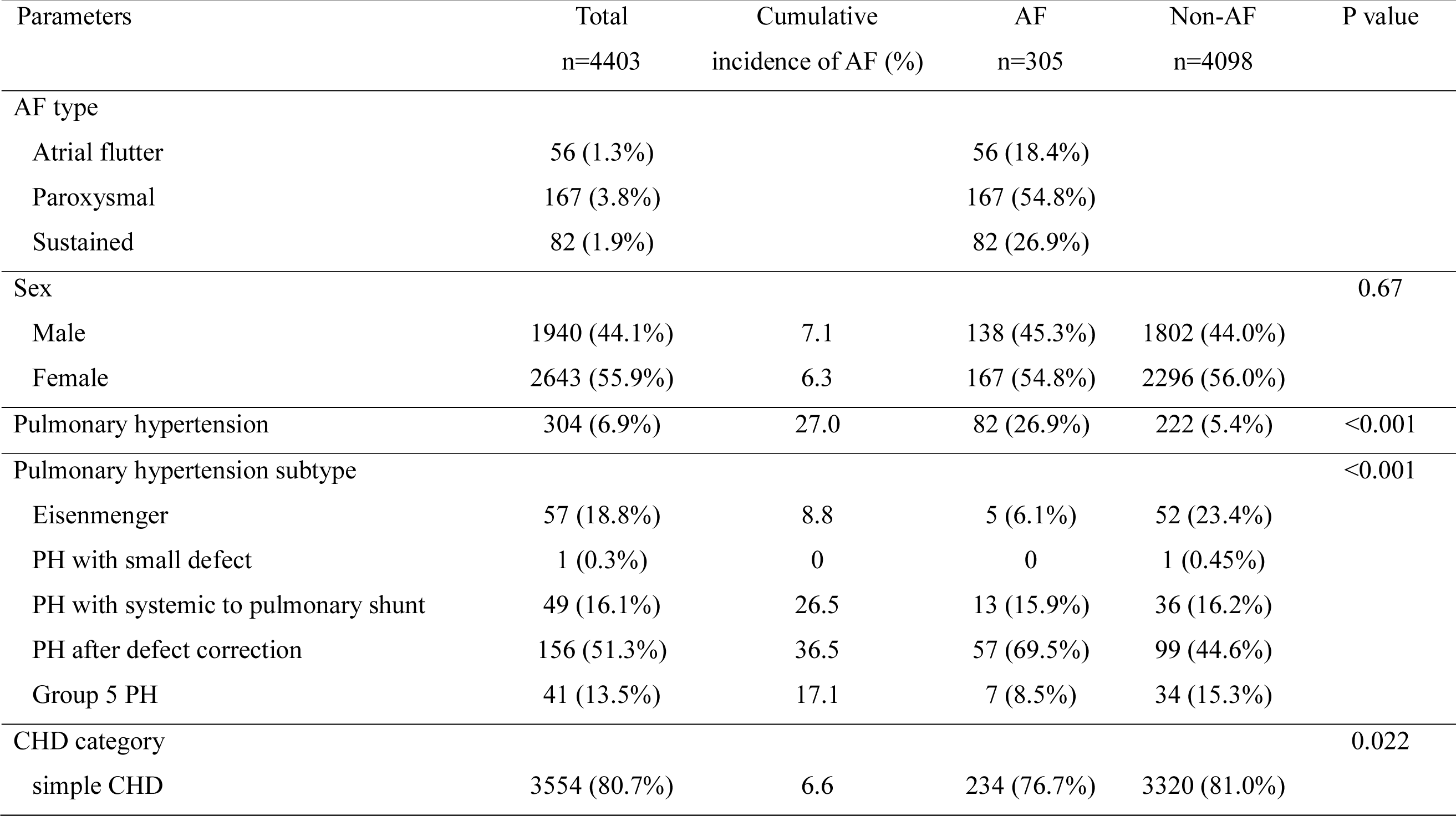
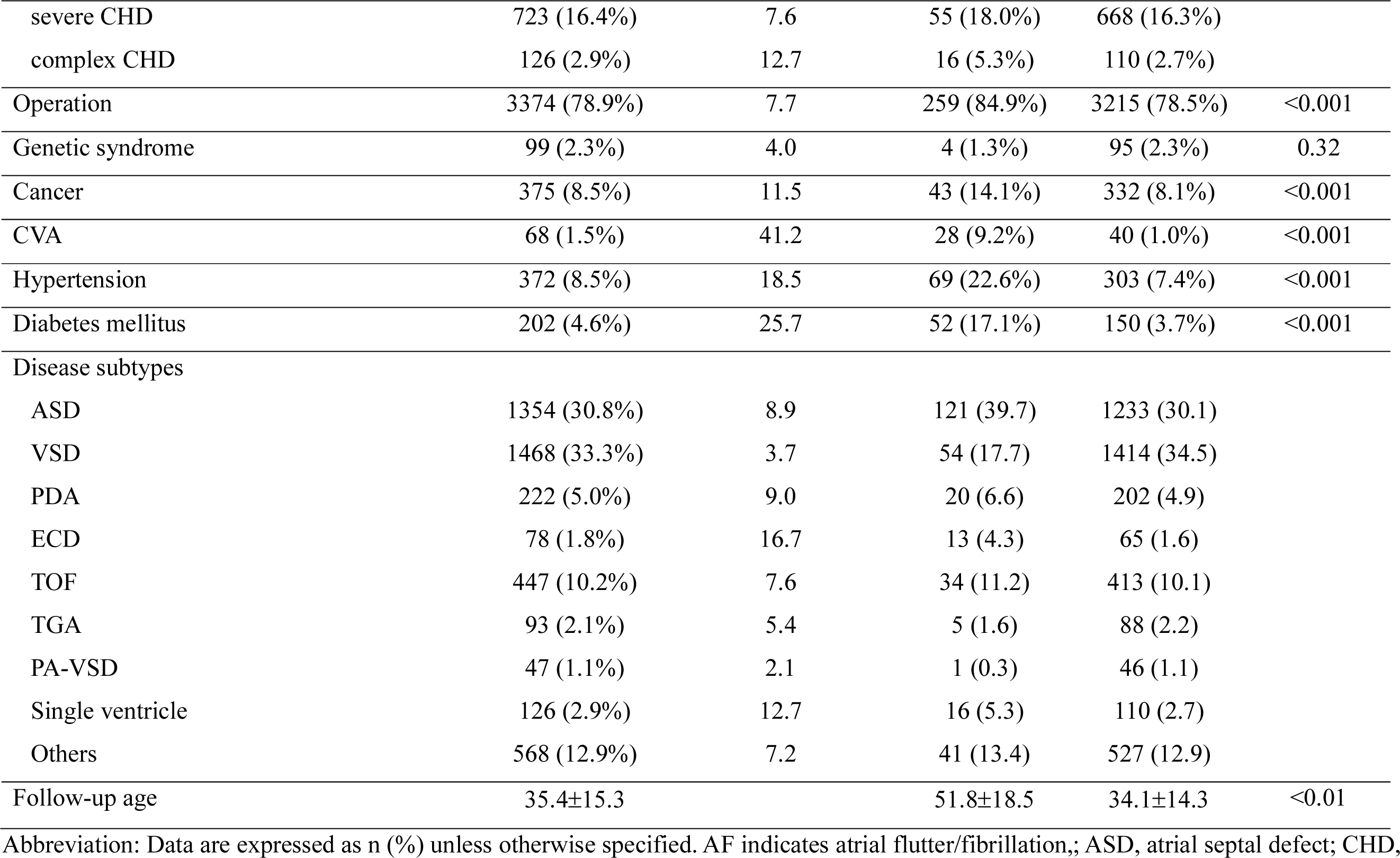
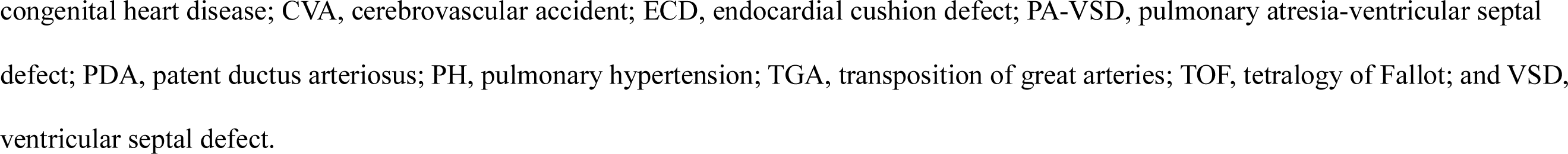
Baseline Demographics and Cumulative Incidence of AF in a Large Cohort with Adult CHD

| Parameters | Total<br>n=4403 | Cumulative<br>incidence of AF (%) | AF<br>n=305 | Non-AF<br>n=4098 | P value |
| --- | --- | --- | --- | --- | --- |
| AF type |  |  |  |  |  |
| Atrial flutter | 56 (1.3%) |  | 56 (18.4%) |  |  |
| Paroxysmal | 167 (3.8%) |  | 167 (54.8%) |  |  |
| Sustained | 82 (1.9%) |  | 82 (26.9%) |  |  |
| Sex |  |  |  |  | 0.67 |
| Male | 1940 (44.1%) | 7.1 | 138 (45.3%) | 1802 (44.0%) |  |
| Female | 2643 (55.9%) | 6.3 | 167 (54.8%) | 2296 (56.0%) |  |
| Pulmonary hypertension | 304 (6.9%) | 27.0 | 82 (26.9%) | 222 (5.4%) | <0.001 |
| Pulmonary hypertension subtype |  |  |  |  | <0.001 |
| Eisenmenger | 57 (18.8%) | 8.8 | 5 (6.1%) | 52 (23.4%) |  |
| PH with small defect | 1 (0.3%) | 0 | 0 | 1 (0.45%) |  |
| PH with systemic to pulmonary shunt | 49 (16.1%) | 26.5 | 13 (15.9%) | 36 (16.2%) |  |
| PH after defect correction | 156 (51.3%) | 36.5 | 57 (69.5%) | 99 (44.6%) |  |
| Group 5 PH | 41 (13.5%) | 17.1 | 7 (8.5%) | 34 (15.3%) |  |
| CHD category |  |  |  |  | 0.022 |
| simple CHD | 3554 (80.7%) | 6.6 | 234 (76.7%) | 3320 (81.0%) |  |
| severe CHD | 723 (16.4%) | 7.6 | 55 (18.0%) | 668 (16.3%) |  |
| complex CHD | 126 (2.9%) | 12.7 | 16 (5.3%) | 110 (2.7%) |  |
| Operation | 3374 (78.9%) | 7.7 | 259 (84.9%) | 3215 (78.5%) | <0.001 |
| Genetic syndrome | 99 (2.3%) | 4.0 | 4 (1.3%) | 95 (2.3%) | 0.32 |
| Cancer | 375 (8.5%) | 11.5 | 43 (14.1%) | 332 (8.1%) | <0.001 |
| CVA | 68 (1.5%) | 41.2 | 28 (9.2%) | 40 (1.0%) | <0.001 |
| Hypertension | 372 (8.5%) | 18.5 | 69 (22.6%) | 303 (7.4%) | <0.001 |
| Diabetes mellitus | 202 (4.6%) | 25.7 | 52 (17.1%) | 150 (3.7%) | <0.001 |
| Disease subtypes |  |  |  |  |  |
| ASD | 1354 (30.8%) | 8.9 | 121 (39.7) | 1233 (30.1) |  |
| VSD | 1468 (33.3%) | 3.7 | 54 (17.7) | 1414 (34.5) |  |
| PDA | 222 (5.0%) | 9.0 | 20 (6.6) | 202 (4.9) |  |
| ECD | 78 (1.8%) | 16.7 | 13 (4.3) | 65 (1.6) |  |
| TOF | 447 (10.2%) | 7.6 | 34 (11.2) | 413 (10.1) |  |
| TGA | 93 (2.1%) | 5.4 | 5 (1.6) | 88 (2.2) |  |
| PA-VSD | 47 (1.1%) | 2.1 | 1 (0.3) | 46 (1.1) |  |
| Single ventricle | 126 (2.9%) | 12.7 | 16 (5.3) | 110 (2.7) |  |
| Others | 568 (12.9%) | 7.2 | 41 (13.4) | 527 (12.9) |  |
| Follow-up age | 35.4±15.3 |  | 51.8±18.5 | 34.1±14.3 | <0.01 |

Abbreviation: Data are expressed as n (%) unless otherwise specified. AF indicates atrial flutter/fibrillation;; ASD, atrial septal defect; CHD,
congenital heart disease; CVA, cerebrovascular accident; ECD, endocardial cushion defect; PA-VSD, pulmonary atresia-ventricular septal defect; PDA, patent ductus arteriosus; PH, pulmonary hypertension; TGA, transposition of great arteries; TOF, tetralogy of Fallot; and VSD, ventricular septal defect.

### Cumulative incidence and risk factors for AF in ACHD patients

Detailed information on the cumulative incidence of AF is presented in Table 1. Of the 4,403 patients enrolled, IART accounted for 18.4% of AF cases, while paroxysmal AF and sustained AF accounted for 54.8% and 26.9%, respectively. The mean onset ages of IART, paroxysmal AF, and sustained AF were 35.7±15.8, 48.4±19.3, and 56.9±14.2 years, respectively.

The highest incidence of AF was seen in those with complex CHD (12.7%), and the onset age of AF was lowest in those with complex CHD, followed by severe CHD (Figure 1A). The AF incidence in specific CHD was shown in figure 2. The AF cumulative incidence increase with age in all types of CHD. By the age of 40, patients of SV and ECD had incidence of AF higher than 30%, and by the age of 60, even simple CHD and severe CHD as TOF has incidence of AF higher than 10%.

**Figure 1:**
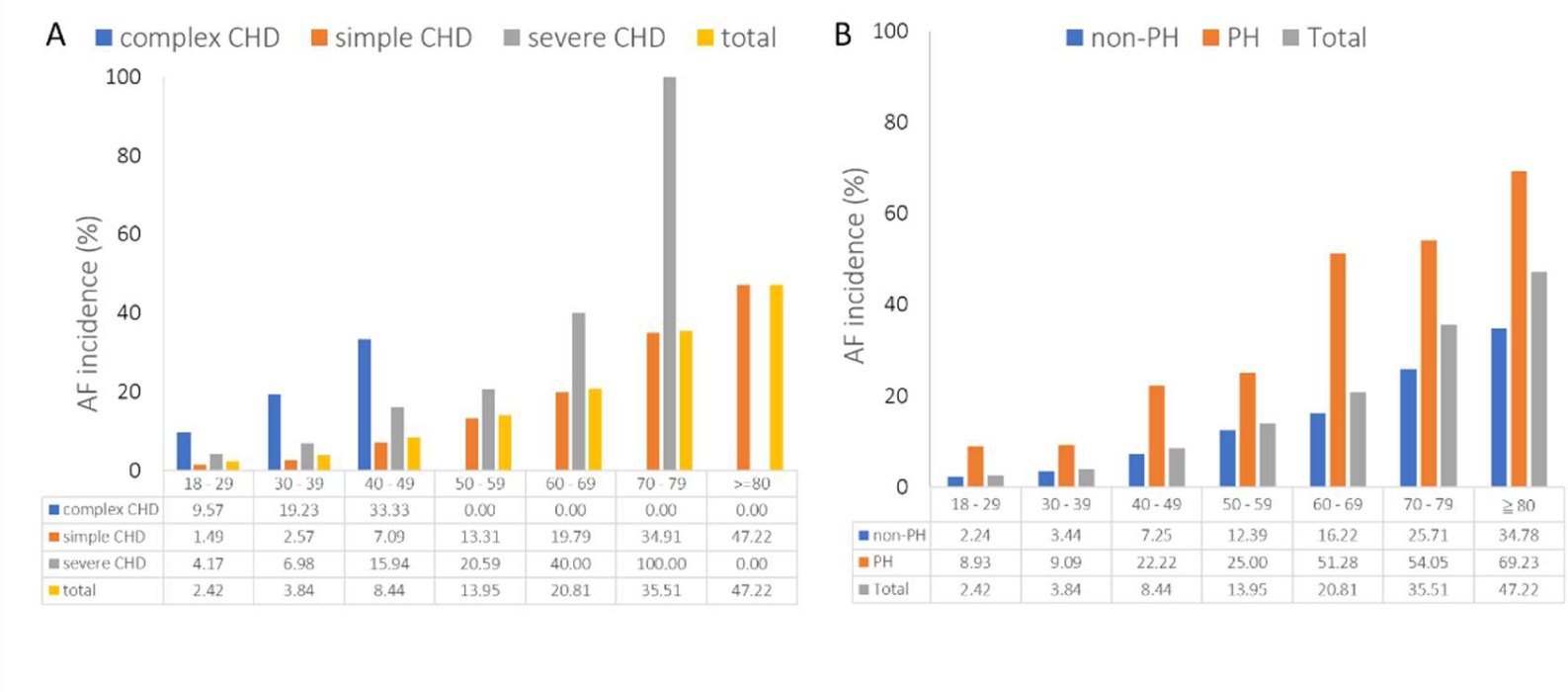
This graph (A) displays the cumulative incidence of atrial fibrillation (AF) by age group in the whole cohort and in different subtypes of congenital heart disease (CHD), as well as the incidence in the presence of pulmonary hypertension (PH) (B). The cumulative incidence rate of AF increases with age in the whole cohort and in each CHD subtype. The incidence rate increases early and is the highest in complex CHD, reaching 33% by age 40. The incidence rate is also higher in severe CHD compared to that of simple CHD. The incidence of AF is higher in the presence of PH and rapidly increased by age 40, reaching 51% by age 60. The incidence rates of specific category in each age group are labelled on the x-axis below the graph.

**Figure 2:**
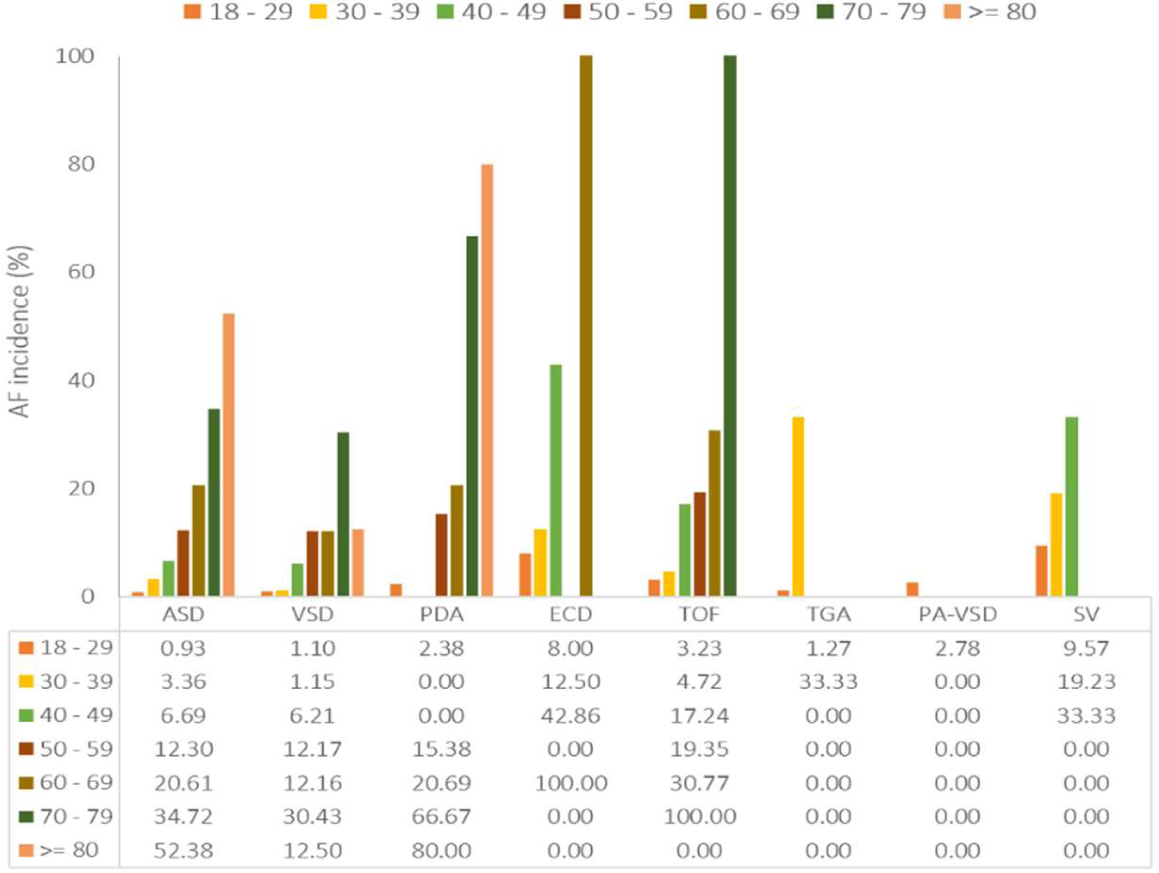
This graph shows the cumulative incidence of atrial fibrillation (AF) in specific congenital heart disease (CHD) by age group. The cumulative incidence rate of AF increases with age in all types of CHD, with the highest incidence in ECD and SV. By the age of 50, the cumulative incidence is higher than 30%. By the age of 60, almost all types of CHD have a cumulative incidence higher than 10%. Abbreviation: ASD, atrial septal defect; VSD, ventricular septal defect; PDA, patent ductus arteriosus; ECD, endocardial cushion defect; TOF, tetralogy of Fallot; TGA, transposition of great arteries; PA-VSD, pulmonary atresia-ventricular septal defect; SV, single ventricle disease.

Patients with PH had a higher risk and earlier onset of AF compared to those without PH (Figure 1B). AF incidence increase with age in both groups. The cumulative incidence rate of AF was 27% in patients with PH, with the highest incidence observed in those with PH after defect correction (36.5%), followed by those with PH and systemic to pulmonary shunt (26.5%).

Patients with a history of previous interventions, metabolic syndrome (diabetes and hypertension), and CVA had higher cumulative incidence rates of AF, with CVA patients having the highest rate of 41.2% (Table 1). Multivariate Cox regression analysis (Table 2) showed that the most significant risk factors for AF development were PH, male sex, severe and complex CHD (odds ratios of 1.91, 1.67, 3.55, and 12.6, respectively).

**Table 2.**
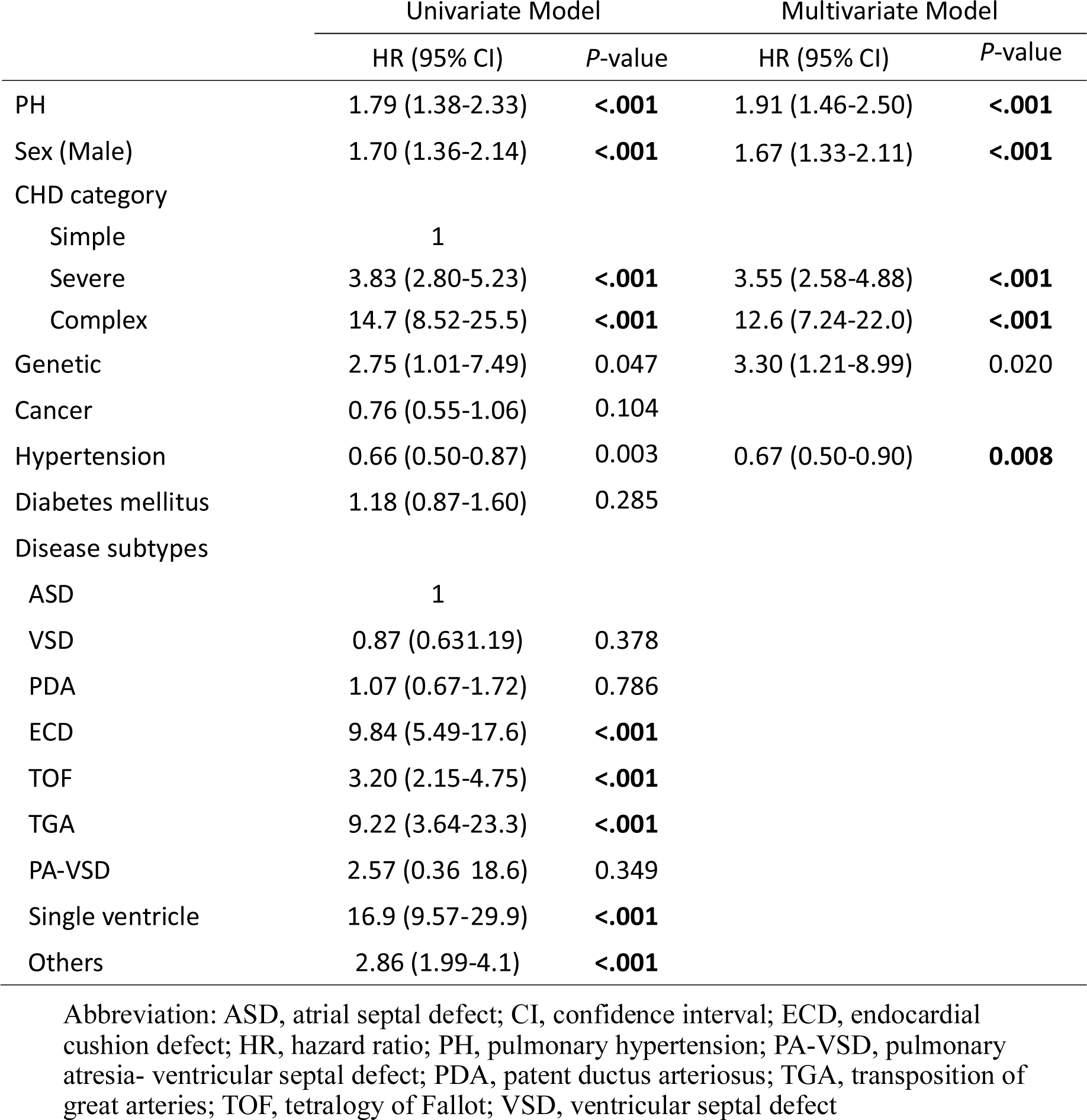
Cox regression analysis for risks of atrial flutter/fibrillation in patients with adult congenital heart disease (ACHD)

|  | Univariate Model |  | Multivariate Model |  |
| --- | --- | --- | --- | --- |
|  | HR (95% CI) | P-value | HR (95% CI) | P-value |
| PH | 1.79 (1.38-2.33) | **<.001** | 1.91 (1.46-2.50) | **<.001** |
| Sex (Male) | 1.70 (1.36-2.14) | **<.001** | 1.67 (1.33-2.11) | **<.001** |
| CHD category |  |  |  |  |
| Simple | 1 |  |  |  |
| Severe | 3.83 (2.80-5.23) | **<.001** | 3.55 (2.58-4.88) | **<.001** |
| Complex | 14.7 (8.52-25.5) | **<.001** | 12.6 (7.24-22.0) | **<.001** |
| Genetic | 2.75 (1.01-7.49) | 0.047 | 3.30 (1.21-8.99) | 0.020 |
| Cancer | 0.76 (0.55-1.06) | 0.104 |  |  |
| Hypertension | 0.66 (0.50-0.87) | 0.003 | 0.67 (0.50-0.90) | **0.008** |
| Diabetes mellitus | 1.18 (0.87-1.60) | 0.285 |  |  |
| Disease subtypes |  |  |  |  |
| ASD | 1 |  |  |  |
| VSD | 0.87 (0.63-1.19) | 0.378 |  |  |
| PDA | 1.07 (0.67-1.72) | 0.786 |  |  |
| ECD | 9.84 (5.49-17.6) | **<.001** |  |  |
| TOF | 3.20 (2.15-4.75) | **<.001** |  |  |
| TGA | 9.22 (3.64-23.3) | **<.001** |  |  |
| PA-VSD | 2.57 (0.36-18.6) | 0.349 |  |  |
| Single ventricle | 16.9 (9.57-29.9) | **<.001** |  |  |
| Others | 2.86 (1.99-4.1) | **<.001** |  |  |

Abbreviation: ASD, atrial septal defect; CI, confidence interval; ECD, endocardial cushion defect; HR, hazard ratio; PH, pulmonary hypertension; PA-VSD, pulmonary atresia- ventricular septal defect; PDA, patent ductus arteriosus; TGA, transposition of great arteries; TOF, tetralogy of Fallot; VSD, ventricular septal defect

### CVA free survival and cause of death in ACHD cohort

After a mean follow-up age of 35.4±15.3, the study found that the CVA free survival rate at ages 40, 50, 60, and 70 was 95.8% (95% CI, 94.9%-96.6%), 92.7% (95% CI, 91.3%-94.0%), 88.4% (95% CI, 86.3%-90.3%), and 77.0% (95% CI, 72.9%-80.9%) respectively. The CVA free survival rate was inversely associated with CHD severity (Fig 3A), with a rate of 44.8%, 92.8%, and 97.3% at age 40 in patients with complex, severe, and simple CHD, respectively (p-value <0.001). The study also found that the CVA free survival rate was lower in patients with AF or PH (log-rank test, p-value <0.001, Fig 3B&C). Multivariate Cox regression analysis identified PH, male sex, severe and complex CHD, and genetic syndrome as the most significant risk factors of primary endpoint (Table 3). Although the presence of AF was not associated with an increased risk of primary outcome after adjusting for other factors, regression analysis showed that AF was independently associated with an increased risk of CVA (Supplementary Table 2). Further analysis showed only paroxysmal or sustained AF but not IART was associated with increased risk of CVA.

**Figure 3:**
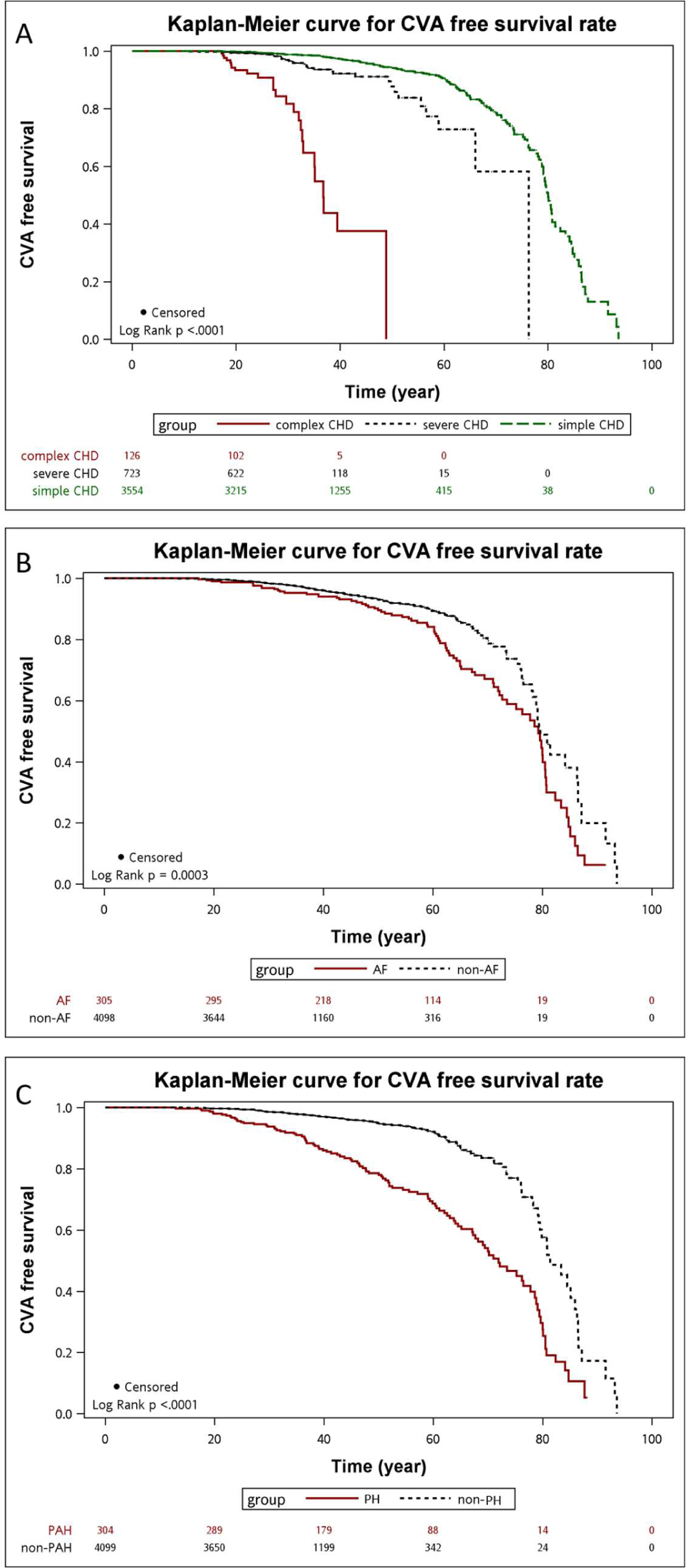
This graph (A) shows the Kaplan-Meier survival curve for cerebrovascular accident (CVA)-free survival in patients with different subtypes of congenital heart disease (CHD), as well as in the presence or absence of atrial fibrillation (AF) (B) and pulmonary hypertension (PH) (C). The CVA-free survival rate iss the lowest in complex CHD and the highest in simple CHD (p<0.01 by log rank analysis). A rapid decline in the CVA-free survival rate is noted at age 25 in complex CHD and at age 50 in severe CHD. The CVA-free survival rate is lower in patients with AF, with a rapid decline by age 60 compared to those without AF. For patients with PH, the CVA-free survival rate is significantly lower compared to those without PH as early as age 20.

**Table 3.** Cox regression analysis for risks of primary endpoint (stroke or mortality) in patients with adult congenital heart disease (ACHD)

|  | Univariate Model |  | Multivariate Model |  |
| --- | --- | --- | --- | --- |
|  | HR (95% CI) | P-value | HR (95% CI) | P-value |
| AF | 1.67 (1.26-2.21) | **<.001** | 1.00 (0.74-1.34) | 0.967 |
| PH | 3.43 (2.64-4.45) | **<.001** | 3.38 (2.54-4.50) | **<.001** |
| Sex (Male) | 1.56 (0.93-2.61) | 0.093 | 1.76 (1.36- 2.29) | **<.001** |
| CHD category |  |  |  |  |
| Simple | 1 |  | 1 |  |
| Severe | 2.66 (1.80-3.92) | **<.001** | 2.62 (1.76-3.89) | **<.001** |
| Complex | 25.0 (15.7-39.8) | **<.001** | 19.3 (11.9-31.4) | **<.001** |
| Genetic | 7.35 (3.70-14.6) | **<.001** | 8.91 (4.43-17.9) | **<.001** |
| Cancer | 1.11 (0.80-1.53) | 0.538 |  |  |
| Hypertension | 0.68 (0.50-0.92) | 0.013 | 0.69 (0.50-0.96) | 0.029 |
| Diabetes mellitus | 1.21 (0.87-1.69) | 0.259 |  |  |
| Disease subtypes |  |  |  |  |
| ASD | 1 |  |  |  |
| VSD | 1.34 (0.97-1.87) | 0.080 |  |  |
| PDA | 1.49 (0.93-2.41) | 0.100 |  |  |
| ECD | 5.74 (2.49-13.2) | **<.0001** |  |  |
| TOF | 2.31 (1.37-3.90) | **0.002** |  |  |
| TGA | 13.0 (5.08-33.3) | **<.0001** |  |  |
| PA-VSD | 6.97 (1.69-28.8) | **0.007** |  |  |
| Single ventricle | 6.97 (1.69-28.8) | **<.0001** |  |  |
| Others | 2.68 (1.75-4.11) | **<.0001** |  |  |

Abbreviation: AF, atrial flutter/fibrillation; ASD, atrial septal defect; CI, confidence interval; ECD, endocardial cushion defect; HR, hazard ratio; PH, pulmonary hypertension; PA-VSD, pulmonary atresia- ventricular septal defect; PDA, patent ductus arteriosus; TGA, transposition of great arteries; TOF, tetralogy of Fallot; VSD, ventricular septal defect

Table 4 showed that heart reasons were the most common cause of death in the entire cohort, both in the AF and non-AF groups. CVA was a more frequent cause of death in the AF group, while cancer and accidents were more common causes of death in the non-AF group.

**Table 4.** Causes of death overall and with/without AF (atrial fibrillation)

| Cause of death | Overall<br>(N=208) | AF (+)<br>(n=64) | AF (-)<br>(n=144) |
| --- | --- | --- | --- |
| Cardiac | 118 (56.73) | 39 (60.94) | 79 (54.86) |
| Cancer | 25 (12.02) | 3 (4.69) | 22 (15.28)* |
| Accident | 12 (5.77) | 0 (0.00) | 12 (8.33)* |
| CVA | 12 (5.77) | 7 (10.94)* | 5 (3.47) |
| Pneumonia | 5 (2.40) | 3 (4.69) | 2 (1.39) |
| Diabetes mellitus | 9 (4.32) | 2 (3.13) | 7 (4.86) |
| Renal | 5 (2.40) | 1 (1.56) | 4 (2.78) |
| Others | 22 (10.57) | 9 (14.06) | 13 (9.03) |

Abbreviation: CVA- cerebrovascular accident
\* $p < .05$
Data expressed as n (%)

## Discussion

From this large Asian cohort study, we have several important findings. Firstly, the cumulative incidence of AF in the entire ACHD cohort was 6.9%. Notably, patients with PH and complex CHD had a higher incidence of AF, and the risk of AF increased with age. Secondly, the CVA-free survival rate was 88.3% by the age of 60 years, and this rate was negatively associated with PH, complex CHD, and genetic syndromes. While AF (especially paroxysmal and sustained AF) increased the incidence of CVA, it did not affect overall survival after controlling for other confounding factors.

### Long-Term complications and AF in ACHD Patients

Patients with ACHD have been shown to have a higher morbidity and mortality rate than the general population. In fact, a Swedish National registry found the mortality rate to be 3.2 times higher than that of the general population^15^. Another study from the Brompton Center reported a mortality rate of 7.3/1000 patient-years^16^. However, with the improvement of congenital heart care, long-term outcomes have improved significantly. The CONCOR registry report found that the median survival for severe, moderate, and simple CHD in the ACHD population was 53, 75, and 84 years, respectively^17^. In a recent report, we found that 95.3% of ACHD patients may survive up to 50 years of age^9^. Nonetheless, long-term complications such as heart failure, stroke, arrhythmia, and PH have gained more attention^18^.

AF is a significant arrhythmia in patients with ACHD, and is a major contributor to morbidity and mortality.^2, 19^. In a multicenter study conducted by Labombarda et al. in North American centers, AF was the second most common presenting arrhythmia, accounting for 29% of cases, following afterward IART. Nevertheless, AF became the predominant arrhythmia in patients aged 50 or older, highlighting the increasing concern for AF in aging CHD patients. Moreover, the incidence of AF typically progresses from paroxysmal to sustained forms as the disease advances^20^. In our study, we observed that the mean onset age of IART, paroxysmal AF, and sustained AF was 35.7, 48.4, and 56.9 years, respectively, which supports the progressive nature of AF in CHD patients over time.

Our study demonstrated that ACHD patients in our Asian country have a similar incidence rate of atrial fibrillation (AF) to Western populations, with a cumulative incidence of 6.9%. This trend increases to 8.4% by the age of 49 and 14% by the age of 59. These results are comparable to the Swedish nationwide registry study, which reported an incidence rate of 8.3% by the age of 42^3^. In contrast, a recent Korean nationwide health insurance dataset reported a lower incidence rate of 4.3% by the age of 49 and 6.8% by the age of 59^8^. However, this may be due to the potential underestimation of the study as it relied solely on diagnosis coding by health insurance data. Our study utilized ECG and Holter reports to improve diagnostic accuracy. Although dietary and lifestyle pattern possibly contribute to the low incidence of AF in the general population in Asian population, this result may not apply to the ACHD population^7^. In the present study, in ACHD patients, AF generally manifests at a younger age, and the main predictors of AF are hemodynamic problems such as PH and CHD severity, rather than metabolic factors. Therefore, the development of AF in ACHD patients is more closely linked to the cardiac hemodynamic mechanism rather than the conventional cardiovascular risk factors.

This may explain why the incidence rate of AF in our Asian cohort is similar to that of Western populations. While dietary and lifestyle patterns may contribute to the lower incidence of AF in the general Asian population, this result may not apply to ACHD patients

### Risk of AF in ACHD patients-complex CHD and PH

Labombarda et al. identified age, hypertension, and smoking as significant risk factors for AF in patients with CHD^20^. Another study suggested that additional factors such as single ventricle disease, previous intracardiac repair, systemic right ventricle, pulmonary hypertension, pulmonary regurgitation, systemic atrioventricular valve dysfunction, and ventricular dysfunction may also increase the risk of AF in these patients^21^. Our findings support that the incidence of AF in ACHD increases with the severity of CHD and the presence of PH as well as advancing age, and underlying metabolic syndrome (hypertension and diabetes). Notably, the first two factors, i.e., the severity of CHD and the presence of PH, appear to have a greater impact on the development of AF.

Patients with single ventricle disease who have undergone a Fontan operation had a high incidence of atrial arrhythmia due to abnormal hemodynamics, which caused high atrial pressure, progressive atrial dilatation, and fibrosis. These changes, combined with previous surgical scarring, can cause abnormal electrical remodeling and served as a substrate for AF^22^. Our study showed that the risk of AF in patients with single ventricle disease is 12.6 times higher compared to those with simple ACHD. Furthermore, the risk of CVA or death is substantially increased by 19.3 times compared to that of simple ACHD. Because the distinct hemodynamic alterations associated with Fontan operation, AF can result in impaired pulmonary venous blood return, leading to the failure of Fontan circulation. These findings underscore the importance of timely detection and appropriate management of AF in single ventricle after Fontan operation patients. Recently, the new modification utilizing total cavopulmonary circulation may lower the incidence of AF^23^. However, it cannot completely eliminate the risk of AF. Our study only focused on adult patients, most of whom underwent Fontan-type operations in the early era. Therefore, the findings of our study cannot be extrapolated to the latter group. Longer follow-up is needed to evaluate the effectiveness of the new modifications in reducing the risk of AF and CVA/death.

In our study, the cumulative incidence of AF in patients with PH was 27%. PH can cause pressure and volume overload in the right atrium and ventricle (if there is a systemic to pulmonary shunt), leading to atrial fibrosis, slow conduction velocity, and the triggering of inhomogeneous conduction, culminating in AF^24^. Additionally, elevated left atrial pressure in patients with left heart dysfunction can lead to left atrial remodeling, increasing the incidence of AF^25^. These mechanisms likely contribute to the high incidence of AF observed in patients with ACHD and PH. During subgroup analysis in our study, among the various subtypes of PH, the highest cumulative incidence of AF was found in patients with PH after defect correction (36.5%), while patients with Eisenmenger syndrome had the lowest incidence. This finding in consistent with a previous study conducted by Drakopoulou et al, which also reported that patients with Eisenmenger syndrome had a lower incidence of new-onset arrhythmia^26^. This suggests that previous atrial surgery and its associated scarring may interact with the pressure and volume overload on the right atrium due to PH, thereby increasing the risk AF in patients of ACHD.

### CVA and outcome in AF with ACHD

AF is a well-established risk factor for CVA and mortality in the general population. Studies have shown that AF increase the risk of stroke by 2.2 to 3.3-fold in the community-based and nationwide health insurance data^6–7^. In patients with ACHD, the prevalence of CVA is high with CVA accounting for 2.0% of ACHD patients^27^. The presence of AF or atrial arrhythmia in ACHD patients increases the risk of CVA by 2.2-2.5 times and the risk of death by 1.5-5.5 fold^2, 3^. The current study found that ACHD with AF had a lower CVA-free survival compared to those without AF, with an odds ratio of 1.69, which is consistent with previous studies^2, 3^. However, after adjusting for other risk factors, the presence of AF only increased the risk of CVA but not the primary endpoint. The different results in our study may have multiple causes. Unlike previous studies that did not include PH and complex CHD in the regression model^2, 3^, this study considers these factors, which are important risk factors for both AF and the primary endpoint. As hemodynamic mechanisms are more crucial for primary endpoint in ACHD patients, this diminish the impact of AF on mortality. Additionally, those with significant hemodynamic problems may have shorter life-expectancy and may not present with AF. This highlights the complex interplay of multiple risk factors in ACHD patients and underscores the need for hemodynamic management in addition to AF treatment.

PH and the severity of CHD were identified as not only risks for AF, but also as crucial risk factors for the primary endpoint. PH has been established as a significant negative predictor of mortality in patients with ACHD^9^, with prior research indicating the presence of PH can increase mortality risk by 4 to 15 times^9^. As described, the adverse effect of PH include increased ventricular pressure loading and atrial pressure, which can increase the risk of AF and worsening heart failure symptoms, ultimately contributing to a higher risk of the primary endpoint in these patients.

Therefore, the management of PH is an essential aspect of treating ACHD patients, as it can help to mitigate the risks associated with both AF and the primary endpoint.

Patients with complex CHD, i.e. single ventricle disease after Fontan type operation, are at a particular high risk of thromboembolism due to endothelial dysfunction, abnormal blood flow, and hypercoagulable state resulting from the unique hemodynamic pattern and associated liver dysfunction^28^. Previous studies have reported rates of thromboembolism and stroke as high as 19%, with subclinical stroke detected through magnetic resonance imaging even more common ^28, 29^. The Australia national registry study showed that the free-from-thromboembolism rate 25 years after Fontan operation is 82%^30^. The TACTIC study reported annual incidence rates of thromboembolic events of 0.93% and 1.95% in severe and complex forms of CHD^31^. Our findings are consistent with these results, demonstrating that complex CHD patients had a CVA-free event rate. These findings underscore the importance of anticoagulant therapy for patients with complex CHD.

In patients with ACHD, paroxysmal or sustained AF, but not IART, was found to be the crucial risk factor for CVA. Although advances in electroanatomical mapping and ablation have considerably improved the outcomes of IART ablation in ACHD patients^32^, the efficacy of AF ablation in ACHD remains limited, which may explain the relatively benign nature of IART in this cohort study^33^. Therefore, managing AF in ACHD patients should prioritize the correction of underlying hemodynamics and consider the limited efficacy of AF ablation.

## Conclusion

This large ACHD cohort showed a high cumulative incidence of AF in this Asian cohort, similar to the Western countries. The incidence increased with age, PH, and CHD severity. CVA-free survival was more closely associated with these factors than with AF.

## Data Availability

The raw data will be provided by the corresponding author if required.

## Acknowledgement

The authors acknowledge the staff of the Department of Medical Research for providing clinical data from the National Taiwan University Hospital-Integrated Medical Database, and the statistical assistance provided by the Center of Statistical Consultation and Research in the Department of Medical Research in National Taiwan University Hospital, Taipei, Taiwan.

---

**Supplementary Table 1.**
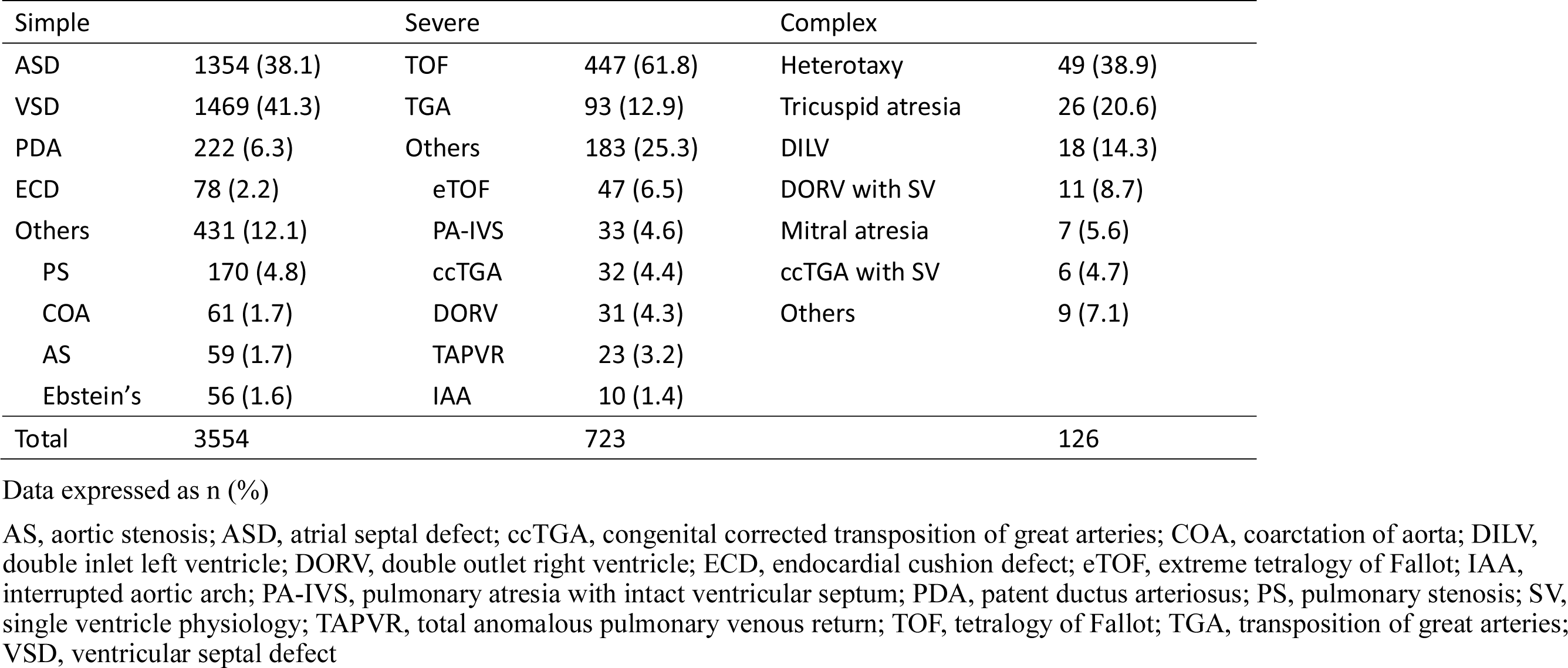
Distribution of congenital heart disease subtypes

**Supplementary table 2.** Cox regression analysis for risks of cardiovascular accident in patients with adult congenital heart disease (ACHD)

|  | Univariate Model |  | Multivariate Model |  |
| --- | --- | --- | --- | --- |
|  | HR (95% CI) | P-value | HR (95% CI) | P-value |
| AF | 2.99 (1.78-5.02) | **<.001** | 2.16 (1.24-3.78) | **0.007** |
| PH | 2.89 (1.72-4.86) | **<.001** | 1.92 (1.09-3.39) | **0.024** |
| Sex (Male) | 1.56 (0.93-2.61) | 0.093 | 1.10 (0.66- 1.83) | 0.728 |
| CHD category |  |  |  |  |
| Simple | 1 |  | 1 |  |
| Severe | 1.64 (0.73-3.71) | 0.233 | 1.85 (1.81-4.23) | 0.146 |
| Complex | 14.6 (5.81-36.7) | **<.001** | 12.6 (4.84-32.8) | **<.001** |
| Genetic | 7.29 (2.20-24.2) | **0.001** | 9.75 (2.87-33.2) | **<.001** |
| Cancer | 1.95 (1.13-3.37) | 0.017 |  |  |
| Hypertension | 1.54 (0.90-2.62) | 0.115 |  |  |
| Diabetes mellitus | 1.63 (0.88-2.99) | 0.118 |  |  |
| Disease subtypes |  |  |  |  |
| ASD | 1 |  |  |  |
| VSD | 0.92 (0.50-1.70) | 0.795 |  |  |
| PDA | 0.4 (0.10-1.69) | 0.214 |  |  |
| ECD |  |  |  |  |
| TOF | 0.88 (0.26-2.93) | 0.833 |  |  |
| TGA | 3.96 (0.51-30.6) | 0.188 |  |  |
| PA-VSD | 6.35 (0.83-48.5) | 0.074 |  |  |
| Single ventricle | 14.4 (5.48-38.0) | **<.0001** |  |  |
| Others | 1.76 (0.80-3.89) | 0.162 |  |  |

Abbreviation: AF, atrial flutter/fibrillation, ASD, atrial septal defect; CI, confidence interval; ECD, endocardial cushion defect; HR, hazard ratio; PH, pulmonary hypertension; PA-VSD, pulmonary atresia- ventricular septal defect; PDA, patent ductus arteriosus; TGA, transposition of great arteries; TOF, tetralogy of Fallot; VSD, ventricular septal defect

## Notes

**Source of Funding:** This work was supported by the National Science and Technology Council in Taiwan (107-2314-B-002-169-MY3 and 110-2314-B-002-086-MY3).

### Competing Interest Statement

The authors have declared no competing interest.

### Clinical Trial

not clinical trial

### Funding Statement

This work was supported by the National Science Council in Taiwan (107-2314-B-002-169-MY3 and 110-2314-B-002-086-MY3).

### Author Declarations

National Taiwan University Hospital Institution Review Board 201806073RINB

